# A Quantitative Systems Pharmacology Model of the Pathophysiology and Treatment of COVID-19 Predicts Optimal Timing of Pharmacological Interventions

**DOI:** 10.1101/2021.12.07.21267277

**Authors:** Rohit Rao, Cynthia J. Musante, Richard Allen

**Affiliations:** Early Clinical Development, Pfizer Worldwide Research, Development and Medical, Cambridge, MA, USA

**Author notes:** **Corresponding Author:** Rohit Rao, Richard Allen.

**Keywords:** COVID-19, Immune response, Viral dynamics, Antivirals, Neutralizing antibodies

## Abstract

A quantitative systems pharmacology (QSP) model of the pathogenesis and treatment of SARS-CoV-2 infection can streamline and accelerate the development of novel medicines to treat COVID-19. Simulation of clinical trials allows *in silico* exploration of the uncertainties of clinical trial design and can rapidly inform their protocols. We previously published a preliminary model of the immune response to SARS-CoV-2 infection. To further our understanding of COVID-19 and treatment we significantly updated the model by matching a curated dataset spanning viral load and immune responses in plasma and lung. We identified a population of parameter sets to generate heterogeneity in pathophysiology and treatment and tested this model against published reports from interventional SARS-CoV-2 targeting Ab and anti-viral trials. Upon generation and selection of a virtual population, we match both the placebo and treated responses in viral load in these trials. We extended the model to predict the rate of hospitalization or death within a population. Via comparison of the *in silico* predictions with clinical data, we hypothesize that the immune response to virus is log-linear over a wide range of viral load. To validate this approach, we show the model matches a published subgroup analysis, sorted by baseline viral load, of patients treated with neutralizing Abs. By simulating intervention at different timepoints post infection, the model predicts efficacy is not sensitive to interventions within five days of symptom onset, but efficacy is dramatically reduced if more than five days pass post-symptom onset prior to treatment.

## Introduction

The coronavirus disease 2019 (COVID-19) pandemic, caused by SARS-CoV-2, a novel coronavirus that emerged in 2019, is a major public health burden worldwide. The pandemic has resulted in more than 200 million confirmed cases and more than 4 million recorded deaths as of September 2021 [1]. While COVID-19 vaccines are highly efficacious [2-5], there is a residual risk of severe cases of COVID-19 for unvaccinated and/or high risk populations [6,7]. Given this residual risk, substantial efforts are being expended to meet this urgent medical need through the development of pharmaceutical interventions such as SARS-CoV-2 neutralizing antibodies and anti-viral therapies [8].

The development of novel pharmaceutical therapeutics, or repurposing of existing therapeutics for COVID-19, is challenging due to the complexity of the disease pathophysiology of viral replication and the associated immune response. This is further compounded by the uncertainties in optimal clinical trial design such as dose (and regimen) selection, inclusion and exclusion criteria, sample collection, and treatment duration. One way to address these challenges is the utilization of quantitative systems pharmacology (QSP) models, which leverage and incorporate existing mechanistic knowledge and data to extrapolate to forward predictions [9,10]. To this end, several within-host systems models of COVID-19 were recently developed to elucidate the relative importance of biological processes underlying COVID-19 pathophysiology and evaluate the efficacy of various therapeutic interventions. These mathematical models provide mechanistic support for a link between the disease severity and the timing of Type-1 interferon (IFN) activation after infection [11], an impaired CD8+ T-cell-dependent adaptive immune response [12], and post-hospitalization viral load dynamics [13]. Furthermore, mechanistic model-based analyses suggest that the efficacy of virus-targeting therapeutic interventions declines sharply with the time of intervention relative to symptom onset, [14-18]. While these models have yielded valuable insights about COVID-19 disease pathophysiology and potential therapeutic strategies in general, there is a need for the development of integrated systems models capable of more quantitatively describing key readouts from emerging interventional randomized-controlled clinical trials (RCTs) in COVID-19 patients to inform and accelerate the development of novel COVID-19 therapeutics.

We previously published a prototype model of the immune response to SARS-CoV-2 infection for collaborative development with the aim of shortening the timeline of the traditional cycle for QSP model development given the rapidly evolving nature of the pandemic [19]. An important aspect of QSP model development involves quantitatively recapitulating the heterogeneity observed in clinical populations through the generation of a robust virtual population [20]. Briefly, our typical strategy is to generate a preliminary set of ‘plausible’ parameter sets, then further refine the ‘plausible population’ into a ‘virtual population’ by comparison to randomized controlled trial data. Here we report an updated version of the model with plausible and virtual populations. The initial plausible population of parameter sets are constrained against a curated dataset of published observational clinical studies in COVID-19 patients, which span viral load and immune responses in plasma and lung. The physiologically constrained plausible population is then mapped onto clinically employed COVID-19 disease severity metrics [21] by using plasma IL-6 levels as a key biomarker correlated with disease severity. The plausible population is finally refined to generate a robust virtual population and partially validated using recently emerging interventional RCTs investigating the efficacy of neutralizing Ab cocktail and anti-viral therapeutics in outpatients with COVID-19 [22-25]. To our knowledge, this is currently the only integrated QSP model of within-host SARS-CoV-2 viral dynamics and the immune response to quantitatively capture key virological and clinical endpoints upon treatment of COVID-19 outpatients in interventional, RCTs through the development of a robust virtual population.

## Methods

### Overview of Model Structure

Broadly, the mathematical model links the within-host viral dynamics of SARS-CoV-2 to the activation of the innate and adaptive immune response and the accumulation of tissue damage as a result of pro-inflammatory mediated cell death. A high-level schematic of the salient interactions accounted for in the model is depicted in **Figure 1**, with a brief description in the following text. A more detailed description of the mechanistic interactions in our model can be found in Dai et al [19].

**Figure 1:**
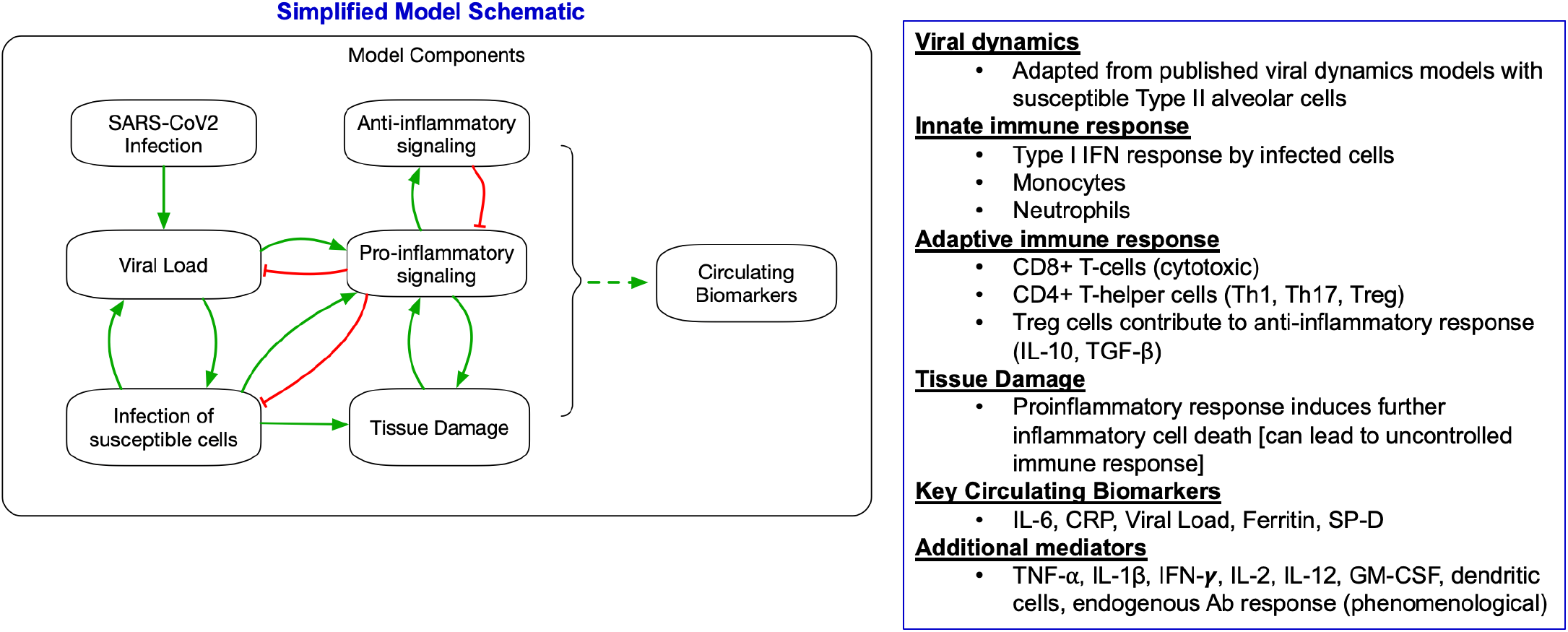
Simplified model schematic: The model describes the productive viral infection of susceptible Type II alveolar cells - infected cells together with free virus activate proinflammatory mediators of the innate and adaptive immune systems (chiefly Type I interferons and CD8+ T-cells) to clear the infected cells. The activation of this proinflammatory response engages anti-inflammatory mediators such as Treg cells, IL-10, and TGF-α, which contribute to resolve the proinflammatory response. Importantly, the proinflammatory response also causes the accumulation of tissue damage as a result of the inflammatory death of infected and bystander alveolar cells. This can lead to positive feedback leading to a sustained immune response indicative of the more severe outcomes of COVID-19. Finally, these processes are linked to certain circulating biomarkers of interest including IL-6, CRP, ferritin, and surfactant protein-D. A more detailed model schematic can be found in Figure 1 in Dai et al. [19].

Briefly, we developed a set of ordinary differential equations (ODEs) to describe the dynamics of SARS-CoV-2 viral load, adapted from previously published models of viral infection dynamics and the innate and adaptive immune responses [26-31]. In our model, uninfected susceptible alveolar Type II cells are infected by SARS-CoV-2 to form productively infected cells, which shed viable virus. The modeled viable virus is representative of clinically measured viral loads upon polymerase-chain reaction (PCR) assay of nasopharyngeal swab samples. Viable virus and infected cells activate the innate and adaptive mediators of the host immune response. Infected alveolar cells produce Type I IFN, which forms an integral part of the innate immune response in our model by preventing the infection of additional susceptible cells and, thus, implicitly accounting for the anti-viral effects of IFN-stimulated gene products [32].

The model further describes the virus and infected cell-induced maturation of macrophages and neutrophils as well as dendritic cells, which are the primary antigen presenting cells (APC) responsible for activating the adaptive immune response. The activated CD8+ cytotoxic T-cells (CTLs) are the key adaptive immune mediators involved in the clearance of infected cells, while the activated CD4+ Th1 and Th17 cells are assumed to maintain a permissive inflammatory milieu through the secretion of proinflammatory cytokines that potentiate the activation of the CTLs. Finally, the model accounts for various clinically relevant biomarkers including proinflammatory cytokines such as IL-6, C-reactive protein, ferritin, and surfactant protein D. For simplicity, the model considers only the alveolar and plasma compartments, corresponding to the major site of infection and the primary site for bioanalytical sample collection, respectively.

The preliminary version of the model [19] accounted for the saturable maturation kinetics (with Hill coefficient =1) of the macrophages, neutrophils and dendritic cells by the virus and infected cells. Importantly, in the current refined model, we hypothesize that in the context of an ongoing infection with an exponentially proliferating pathogen, these cells behave as logarithmic sensors such that the production of mature innate immune cells due to viable virus and infected cells varies in a log-dependent manner [Supplementary materials: Supplementary equations]. This enables the host immune system to remain responsive to pathogen levels ranging over orders of magnitude. Without this characteristic the model can match viral load data but predicts marginal improvements on disease severity following viral load lowering therapies that are administered after the peak in the viral load occurs. This is largely because, with the prior implementation, the immune response is saturated at viral load levels below that observed in treated patients who are observed to have reductions in severity. While still an active area of research, a number of theoretical and experimental studies support the existence of signaling architectures capable of log-sensing (often referred to as the Weber-Fechner property) in varied biological sensory systems, including the immune system [33-37]. Sontag [33] developed a log-sensing network architecture that reproduced prior experimental results by Johansen et al. [38] where exponential increases in antigen stimulation resulted in the greatest immune activation relative to constant or linear antigen stimulation, suggesting that the immune response can detect exponentially increasing pathogen populations due to its lack of adaptation to exponential ramps. More recently, Nienaltowski found that the fraction of the total population of immune cells activated in response to inflammatory cytokine stimuli varied with the logarithm of the stimulus [37]. For simplicity we assume that the log-dependent activation of the immune system is due to activation of the innate immune cells, and not the adaptive immune T-cells. Moreover, we also do not propose specific intracellular or extracellular motifs that give rise to log-sensing given that this is still an active area of research with multiple feasible formulations [34,36].

### Generating a plausible population

We used a tiered approach to calibrate the model and generate a robust virtual population. We first generated an initial plausible population that constrains the model states, such as the viral load and various immune mediators to physiologically reasonable values, such that they qualitatively match a curated collection of observational studies on COVID-19 summarized in Supplementary Table S1. Observational clinical datasets were selected by prioritizing studies with 1) longitudinal measurements of cytokines and immune cell populations in plasma and, where possible, the bronchioalveolar space, 2) longitudinal nasopharyngeal viral load, and 3) other plasma biomarker measurements stratified by disease severity which record the time of measurements relative to symptom onset. When possible, we selected studies with concordant assay sensitivities and read-outs. The observational data used to constrain the model states in the plausible population are primarily from hospitalized COVID-19 patients since such datasets were more readily available and of higher quality at the time of model development; however, we used outpatient data when possible, to inform some of these states such as plasma IL-6 and the viral load [39,40]. Nevertheless, despite the challenges, the data were suitable for the generation of the plausible population given the initial goal of constraining the model to a physiologically realistic regime.

To generate this plausible population, we uniformly sampled biologically relevant parameters [Supplementary Table S2] with high sensitivity and uncertainty within 5-fold bounds of a nominal parameter set and filtered solutions that were within 2-fold bounds of viral load and immune mediator measurements in the curated set of literature data. The incubation period of the infection needs to be assumed to calibrate the time course of simulated infection to the time-dependent dynamics of viral and immune mediators in the curated datasets, which are reported relative to time from symptom onset.

Several epidemiological studies suggest that the viral load of SARS-CoV-2 peaks around the onset of COVID-19 symptoms [13,41-43]. Informed by this epidemiological evidence, we assume that symptom onset coincides with the timing of the peak in viral load for the plausible virtual subjects, thus enabling the translation of the time from ‘day of symptom onset’ to time from ‘day of infection’ in the curated datasets. Thus, rather than assume a fixed, identical SARS-CoV-2 incubation period for each virtual subject, we obtain a distribution of incubation periods across the plausible population based on the individual viral load trajectories of each virtual subject.

Infection with SARS-CoV-2 was simulated using an inoculum equivalent to 10 viral RNA copies/mL. We further account for an endogenous Ab response that begins to have an appreciable effect on viral clearance on Day 20 post infection using a phenomenological representation. While this is roughly in alignment with clinical findings that almost all infected individuals are sero-positive 14-28 days from symptom onset [44], the simplistic phenomenological representation in our model can be more finely refined to these observations as more appropriate clinical datasets become available. Moreover, we assume that, post peak, the virus can no longer infect new susceptible cells within the host when the viral load declines below 10^4^ viral RNA copies/mL. Studies suggest a ratio between 10^3^-10^4^ between PCR assay measurements (in RNA copies/mL) and the number of infectious units measured in tissue culture infective dose (TCID50) [45]. This assumption is made to prevent an unphysiological rebound in viral load at later time points once a low viral load regime (<10^4^ viral RNA copies/mL) post peak is reached.

### Linking to disease severity

The primary outcome in the outpatient interventional RCTs, detailed in subsequent sections, are reported as a reduction of hospitalizations or deaths. We ultimately aimed to generate a final virtual population that would match both the observed reductions in COVID-19 related events in these clinical trials along with the reported changes in viral load markers. Therefore, upon generation of the plausible population, the QSP model outputs must be appropriately translated to clinically reported disease severity categories.

In doing so, we adopt a parsimonious approach and treat plasma IL-6 as the key biomarker correlated to disease severity. Numerous clinical studies provide evidence for a link between increasing plasma IL-6 levels and COVID-19 disease severity and prognosis [46-49]. Del Valle et al., in a large study of > 1400 COVID-19 patients, found a threshold of 70 pg/mL of plasma IL-6 levels at hospitalization could independently predict disease severity and mortality. A recent, RCT evaluating the efficacy of Tocilizumab in >300 subjects further showed plasma IL-6 levels were significantly correlated with baseline clinical severity and were also a significant predictive biomarker for clinical severity through Day 28 of the trial [46,47]. To translate the plausible population to the incidence of hospitalization reported in outpatient clinical trials, a threshold of 40 pg/mL is employed for plasma IL-6 levels, corresponding to a value closer to the lower tail of the plasma IL-6 distribution observed in hospitalized patients with mild-moderate COVID-19 severity [46,47]. Moreover, given the uncertainty in using a plasma biomarker as an indicator of a hard-end point such as hospitalization, we conservatively apply this threshold to the peak IL-6 levels attained at any point over the entire time-course of the simulated infection. Note that this approach is more suitable for estimating event rates in trial populations than the timing of hospitalization or death in specific patients.

### Virtual population refinement to match interventional data

The final virtual population is formed by selecting a subset of plausible subjects whose simulated responses were constrained to interventional data from published RCTs in outpatient COVID-19 patients. Three RCTs were selected for model calibration and validation, specifically, the Blaze-1 Ph3 nAb trial of bamlanivimab and etesevimab (NCT04427501) [22], the Ph2 and Ph3 REGN-COV nAb trial of casirivimab and imdevimab (NCT04425629) [24,50], and the Ph2 interim analysis of the anti-viral molnupiravir [MK-4422/EIDD-2801] (NCT04405570) [25]. These three RCTs primarily evaluated the effectiveness of their respective pharmaceutical interventions in reducing the rate of hospitalization in outpatients with mild to moderate COVID-19 at high risk of hospitalization.

The above RCTs reported the mean viral load dynamics, with the nAb trials further reporting the rate of medically related events through Day 29 of the trial, in the placebo and treatment arms, respectively and the associated relative risk reduction in event rate upon treatment. Thus, a final virtual population was selected from the plausible population such that it matched both the reported viral load time course and the disease severity rates in the placebo and treatment arms of the trial using importance sampling methods previously published in Allen et al. [51]. The virtual population matching the Blaze-1 clinical trial observations was selected such that it was of comparable in size to the trial population (N=516). Since the clinically observed viral load time-course is reported relative to the start of treatment, it was necessary for us to assume a time of infection to calibrate the QSP model. As for the plausible subjects, the time of the peak viral load post simulated infection was assumed to coincide with the time of symptom onset. In the case of each of the simulated interventions, the time of intervention relative to the simulated time of symptom onset (time to peak viral load) is given by the mean time from symptom onset to randomization reported for the corresponding clinical trial.

Despite generating the final virtual population by calibrating the model to only the viral load measurements and severity information available in the interventional trials, our tiered model calibration approach, described above, ensures all other model states are still constrained to physiologically plausible values informed by the curated set of observational clinical data in COVID-19 patients.

#### Modeling neutralizing antibody therapeutics

The pharmacodynamic effect of the nAb cocktails are modeled to decrease the rate constant for the production of infected cells due to viable virus. This is informed by their mechanism of action whereby the nAbs selectively bind to the spike protein of SARS-CoV-2, thus neutralizing the virus particles, preventing their entry into susceptible cells, and subsequent replication [52].

##### Blaze-1 Ph3 nAb trial

The efficacy of the nAb cocktail bamlanivimab and etesevimab was evaluated in an RCT for outpatients with recently diagnosed, mild to moderate COVID-19. A two-compartment model was used to describe the plasma pharmacokinetics of bamlanivimab and etesevimab. The model parameters and equations were adapted from the publicly available Emergency Use Authorization (EUA) document for the nAb cocktail [Supplementary materials: Supplementary equations] [23]. The QSP model was calibrated to the Blaze-1 Ph3 placebo and 2800mg bamlanivimab and 2800mg etesevimab treatment arms. The maximal effect (Emax) and the potency (EC50) of the individual nAbs was informed by the pre-clinical and clinical values reported in the EUA document and further optimized to match the observed viral load time course and severity improvements from the clinical trial.

##### REGEN-COV nAb trials

The pharmacokinetics of REGEN-COV (casirivimab and imdevimab) were described using a one-compartment model matched to produce the reported noncompartmental analysis (NCA) parameters from [24], including the maximal plasma concentration (C_max_), plasma concentration at Day 28 post administration (C_Day 28_), and half-life for each antibody, respectively [Supplementary Figure S1]. The QSP model was matched to the placebo and 8g REGEN-COV treatment arms from the REGEN-COV2 Ph2 trial. As partial validation of the simulated Blaze-1 placebo group time-course, a subset of virtual subjects from the virtual population was sampled such that their selection was informed only by the baseline placebo viral load measurement for each of the reported subgroups from the REGEN-COV Ph2 trial. Subsequently, the predicted viral load time-course of the virtual population subset was validated against the entire viral load trajectories of the subgroup placebo arm viral load time courses from this trial.

The maximal effect (Emax) was fitted to match the Ph2 trial observations. The potency (EC50) of the individual nAbs was informed by the published pre-clinical in vitro estimates [52] and further optimized to match the observed viral load time course and severity improvements from the Ph2 clinical trial. The simulated treatment dynamics of the REGEN-COV Ph2 trial, and the suitability of our log-sensing hypothesis were further validated against the lower dose 2.4mg REGEN-COV treatment arm from the Ph3 trial.

#### Modeling anti-viral therapeutics

The anti-viral, molnupiravir is the pro-drug of the pharmacologically active EIDD-1931, a nucleoside analogue which acts by introducing random point mutations throughout the SARS-CoV-2 viral RNA, leading to error catastrophe of viable virus [53]. Informed by this mechanism of action, the pharmacodynamic effects of molnupiravir are modeled as inhibiting the production of viable virus from infected cells. The plasma concentration of EIDD-1931 for model simulations were obtained from published clinical literature [54] [Supplementary Figure S2]. The Emax of the therapeutic is fixed to 1, informed by preclinical in vitro assay information for the active form of the molecule, EIDD-1931 [53]. The EC50 was optimized to match the viral load time course from the Ph2 interim analysis [25]. In modeling the anti-viral molnupiravir, as a simplification we do not attempt to explicitly model the intracellular concentration of its active metabolite EIDD-1931 and instead optimize the EC50 relative to the reported plasma concentration of EIDD-1931.

### Preliminary Virtual Population of SARS-CoV-2 Variants of Concern

The Delta variant is currently the dominant variant of concern worldwide and is potentially associated with an increased risk of post-vaccination breakthrough infection and greater infectiousness [55]. Delta variant SARS-CoV-2 is reported to have a higher peak viral load (lower Ct value), higher viral load upon controlling for days from symptom onset, a longer duration of viral load shedding and a potentially shorted incubation period [56-61]. We selected a subset of virtual subjects from the previously developed plausible population with viral dynamics that are in general agreement with the virological observations from the above-mentioned epidemiological studies. We match data presented in Li et al. [59], describing the greater viral load in Delta variant infections in a cohort of isolated close contacts of individuals with confirmed SARS-CoV-2 infection. Given the Blaze-1 Ph3 clinical trial was completed in early 2021, prior to the emergence of the Delta variant and thus, did not report a significant proportion of Delta variant infections, we assume viral infections in the Blaze-1 population are more representative of the 19A/19B clade reported in Li et al. Furthermore, since Li et al. did not report a time of first test since estimated close contact exposure, we assume the measurement of viral load is made 2 days post infection, where the viral load in the Blaze-1 population corresponds to the reported viral load of the 19A/19B clade.

### Model Simulation

The model was simulated in MATLAB 2019a, and ode15s was used to integrate the model differential equations. The computation time for a single run of the model was on the order of 1 second. The code is available in full at https://github.com/openPfizer/QSP_model_COVID19 and archived in [62].

## Results

The plausible population generated by constraining the simulated viral load and various immune mediators to physiologically reasonable values is shown in **Figure 2** for selected representative model variables. For a more complete depiction of the dynamics, we represent the plausible population time course from the time of infection. The time course of the plausible population relative to the day of symptom onset, as described in the methods is depicted in Supplementary Figure S3. As evidenced by the substantial variability in the plausible virtual population and associated clinical observations, the QSP model captures the significant heterogeneity in viral load and immune markers and is thus able to represent subjects across the spectrum of disease severity, including mild, moderate, and severe COVID-19 patients. Simulated viral inoculation leads to an exponential increase in viral load, which peaks on average ∼5d post infection, followed by a rapid and steady decline. The increasing viral load engages the innate and adaptive immune response, leading to the secretion of several inflammatory cytokines. In agreement with a canonical anti-viral innate immune response [63], the activation of the innate immune mediators leads to an early peak in the time course of Type I IFN, TNF-α and IL-1β secretion, followed by a more delayed increase in IL-6 and IL-10, the chief cytokine assumed to be a biomarker of disease severity and the major counter-regulatory anti-inflammatory cytokine in the QSP model, respectively.

**Figure 2:**
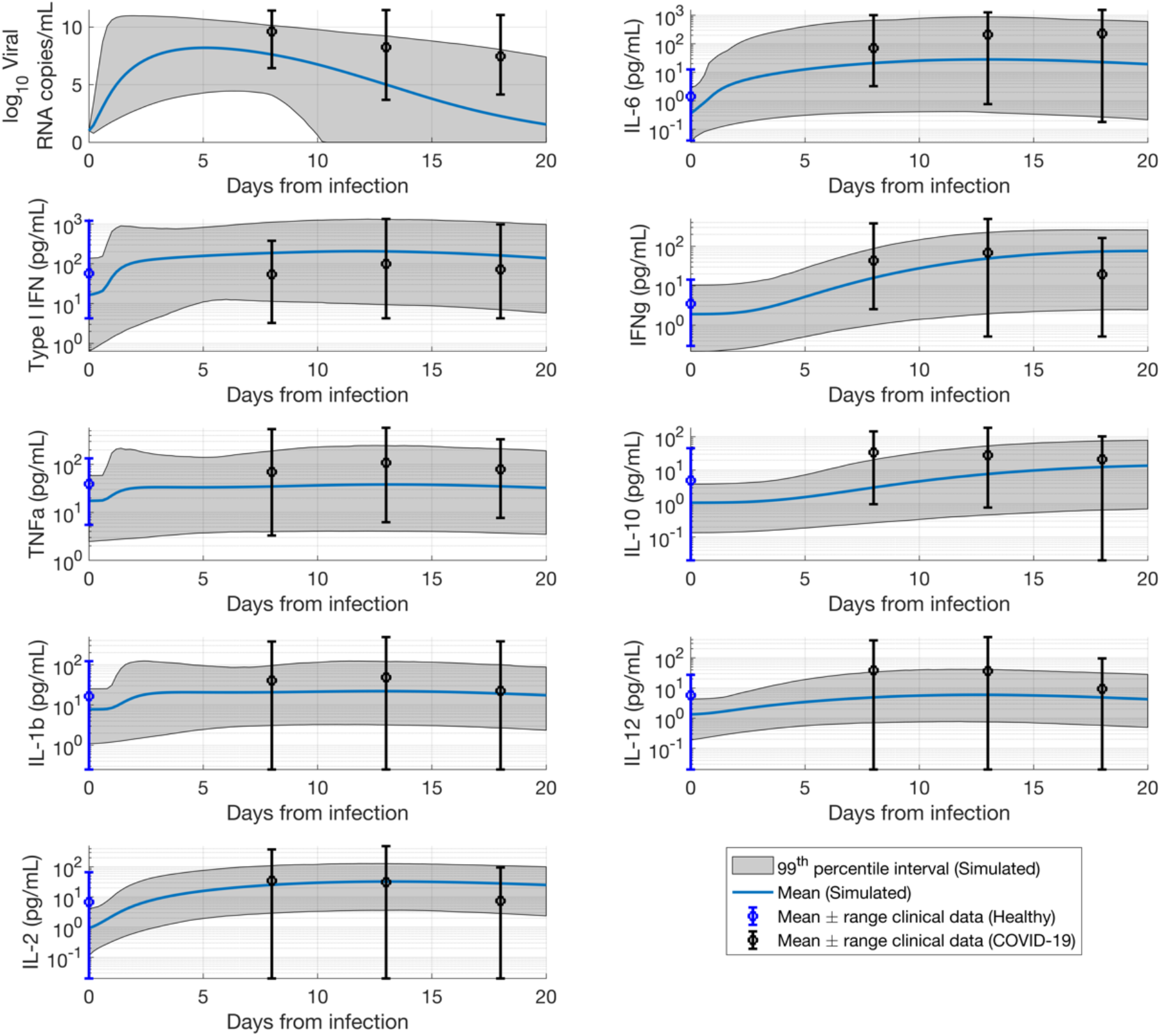
Plausible population overlaid against observational COVID-19 clinical data for the viral load time course and different representative cytokines (N = 14545). For the purpose of visual representation of the time course of the viral and immune makers from the day of infection, the time from symptom onset was translated to a time from infection by assuming an incubation period of 4.5d [64].

Upon generation of the plausible population constrained to the observational clinical studies, a subset of virtual subjects is sampled from the plausible population to form a refined virtual population that quantitatively matches the viral load time course from interventional RCT data. **Figure 3A** depicts the mean simulated viral load time course from the refined virtual population closely matching the time course of the mean trial data from the Blaze-1 Ph3 placebo and 2800mg bamlanivimab and 2800 etesevimab nAb cocktail arms, respectively. While the QSP model adequately captures the more clinically relevant high viral load regime (> 1000 copies/mL), there is a discrepancy between the simulated and clinically observed treatment arms at very low viral loads (below 500 copies/ml) at the later time points of the clinical trial (Day 11). We hypothesize that this discrepancy at low viral loads is due to the SARS-CoV-2 PCR assay characteristics. While the model only accounts for viable virus, the PCR assay can detect inactive, unencapsulated viral fragments [65] that might not be representative of active ongoing infection especially at the low viral loads observed at later time points in the clinical trial. This non-viable viral RNA might persist for much longer than active virus and hence lead to the slower than predicted dynamics (at low viral load) in the time course of the observed treated group. Moreover, this lower viral load regime is also below the reported lower limit of quantification of the PCR assays used in comparable RCTs [24,25].

**Figure 3:**
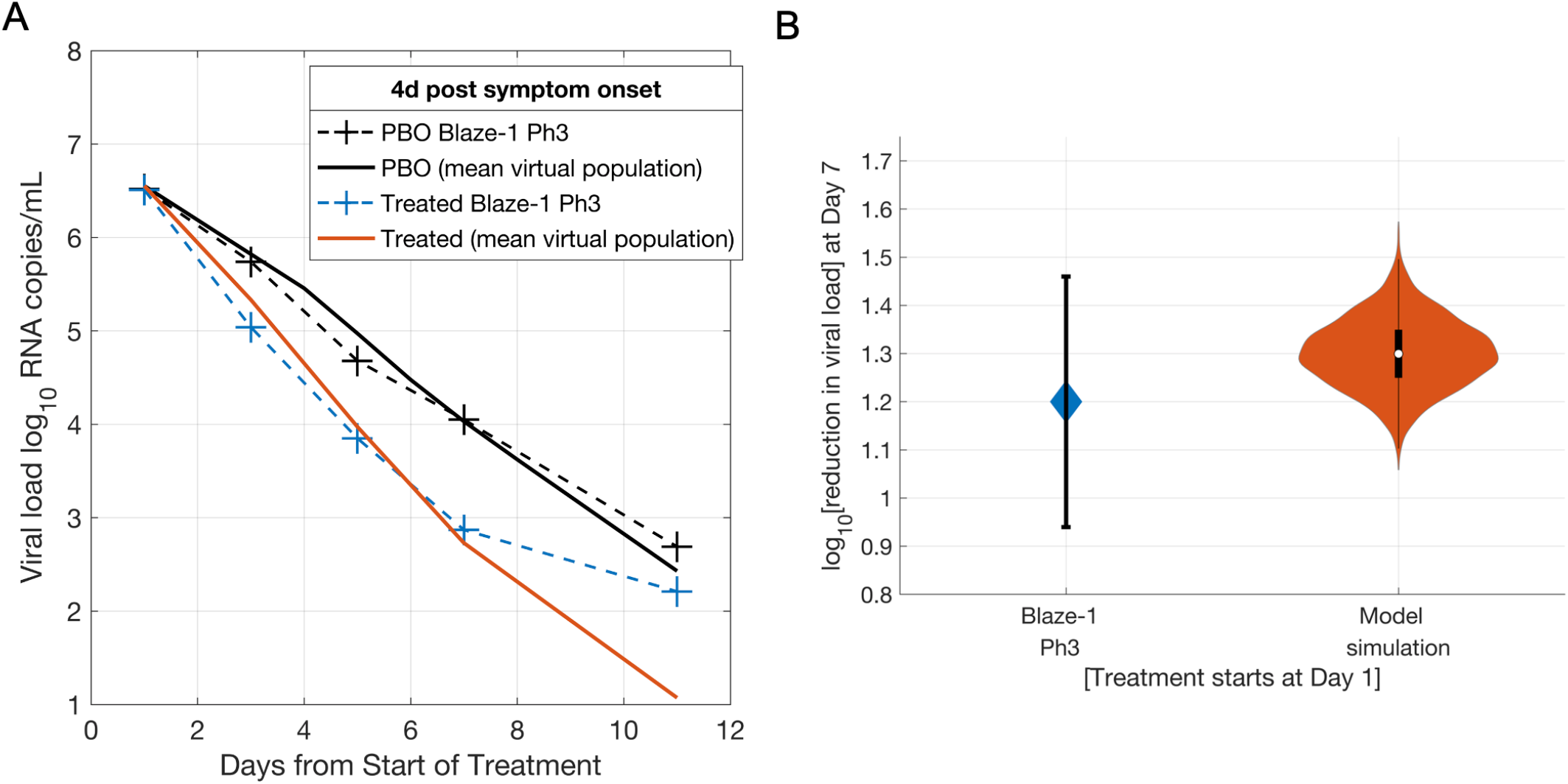
A: Mean of the virtual population (N=502) for the simulated placebo (PBO) group and the 2800mg bamlanivimab + 2800mg etesevimab simulated treated group matching the mean trial data from the observed Blaze-1 Ph3 placebo group and the 2800mg bamlanivimab + 2800mg etesevimab treated group. B: log10 reduction in viral load from baseline at Day 7 after treatment administration in the observed Blaze-1Ph 3 trial (blue) and the simulated the 2800mg bamlanivimab + 2800mg etesevimab treated group. Error bars represent the 95% confidence interval in the mean for clinical trial observations and range of observations in the simulated virtual population. Violin plots are indicative of 99% prediction interval of mean from 1000 bootstrapped samples of the virtual population.

**Figure 3B** depicts both the simulated mean viral load reduction at Day 7 of the clinical trial along with its simulated variability characterized using 1000 bootstrapped trials in good agreement with the corresponding trial observation. The size of the virtual population (N=502) is comparable to that reported in the placebo (N=517) and treatment (N=518) arms. Additionally, the 5.1d mean incubation period for this virtual population, given the assumption that symptom onset occurs at peak viral load, is in good agreement with current epidemiological estimates of the mean incubation period of 4-5d for SARS-CoV-2 [64,66,67]. Moreover, the assumption that intervention occurs post viral load peak in our population is further supported by the observed mean placebo and treatment arm viral load trajectories, which exhibit a monotonic decline from the start of treatment, suggesting that the majority of mild/moderate COVID-19 outpatients enter the trial after the peak in viral load. While we use a threshold of 10^4^ copies/mL to represent an active infection in the current plausible population and Blaze-1 virtual population, our results are not materially affected by a lower threshold of 10^3^ RNA copies/mL, which corresponds to the lower limit of quantification for SARS-CoV-2 PCR assays used in several clinical trials [25,50][Supplementary Figure S4].

Subsequently, the placebo group viral load dynamics of the refined Blaze-1 trial virtual population were partially validated against that from the REGEN-COV Ph2 trial. In addition to an analysis of the overall viral load lowering response upon treatment, the REGEN-COV Ph2 trial also reported a subgroup analysis where subjects were stratified by increasing baseline viral load (i.e. subjects with baseline viral load >10^4^, >10^5^, >10^6^ or >10^7^, respectively). We tested the ability of the model to reproduce the placebo group viral load time course from the REGEN-COV Ph2 trial by using only the baseline viral load measurements for each of the subgroups to inform the selection of a subset of virtual subjects from the Blaze-1 virtual population. **Figure 4A-D** shows the simulated placebo group trajectories from the REGEN-COV virtual population (N=402) in good agreement with the observed placebo group dynamics for each reported subgroup from the Ph2 trial. The model also adequately captures the treatment group trajectories for each subgroup upon fitting the Emax and using preclinical estimates to inform the neutralization EC50 of the simulated REGEN-COV antibodies.

**Figure 4:**
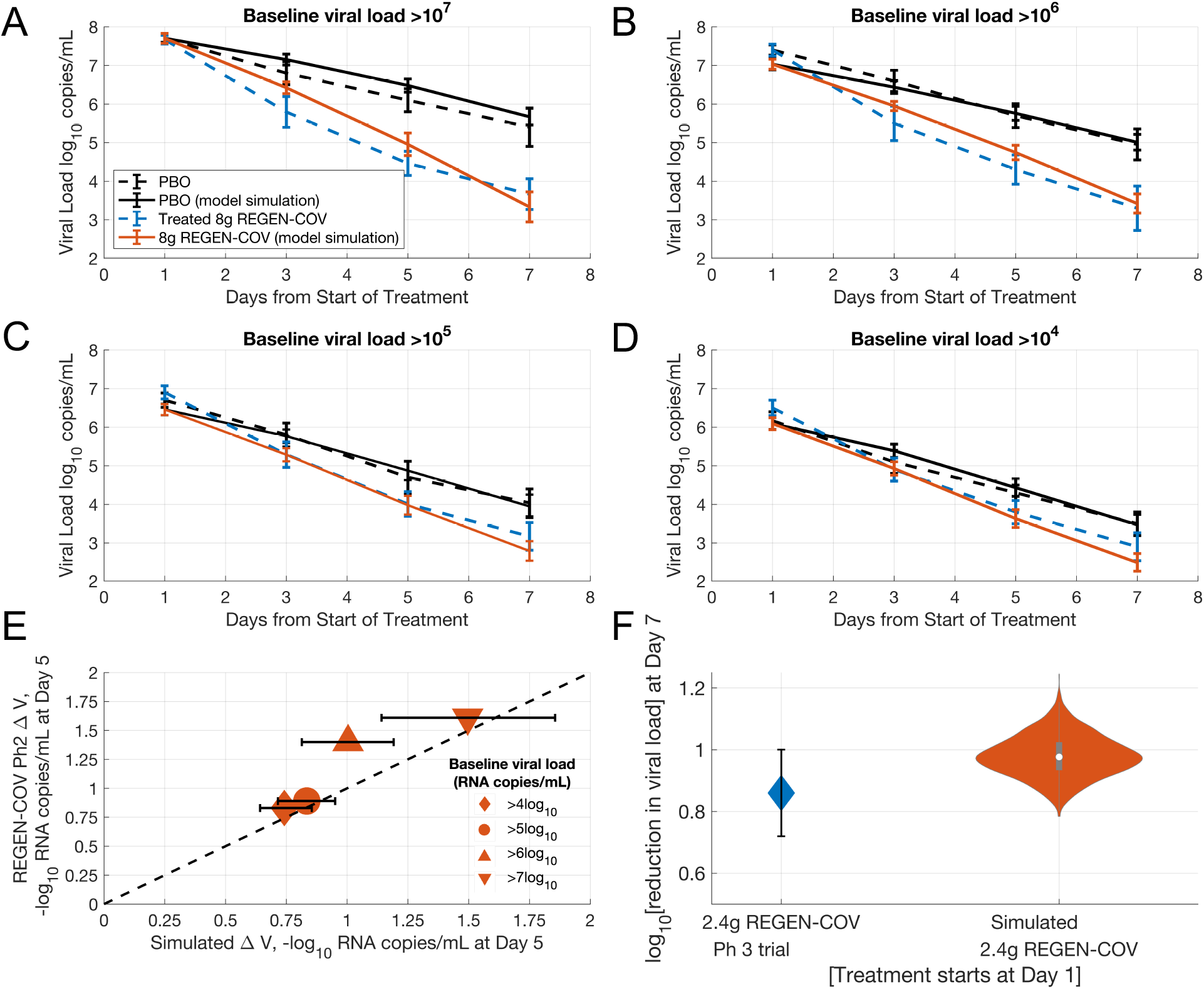
Partial validation of the model against REGEN-COV antibody cocktail trial data: A-D) Time course of the viral dynamics of the overall virtual population and each of the subgroups compared against observations from the REGEN-COV Ph2 clinical trial for the placebo group and the 8g REGEN COV treatment group. Error bars are representative of the standard error for the virtual population and clinical trial observations. E) log10 reduction in viral load from baseline at Day 5 for the overall virtual population and each of the subgroups compared against observations from the REGEN-COV Ph2 clinical trial for the placebo group and the 8g REGEN COV treatment group. Error bars are representative of the 99% prediction interval of the mean for the virtual population F) log10 reduction in viral load from baseline at Day 5 for the overall virtual population compared against observations from the Ph3 trial REGEN-COV trial for the 2.4g REGEN-COV treatment. Error bars are representative of the 95% confidence interval for clinical trial observations. Violin plots are indicative of 99% prediction interval of mean from 1000 bootstrapped samples of the virtual population. REGEN-COV Ph2 data extracted from [24].

Following from this, the model predicts the subgroup responses in agreement with REGEN-COV Ph2 trial findings, where patients with higher baseline viral loads exhibited higher viral load decreases upon treatment [**Figure 4E**]. While the model slightly underpredicts the reduction in viral load for the subgroup with baseline viral load > 10^6^ copies/mL; it is capable of accurately predicting the other subgroup responses and matches this group at day 7. As a further partial validation, using the same virtual population shown in **Figure 4A-D**, and the same pharmacodynamic parameters for REGEN-COV, the model also adequately predicts the viral load lowering efficacy at Day 7 reported in the Ph 3 trial upon simulation of a lower 2.4g REGEN-COV dose [**Figure 4F**].

The model also recapitulates the viral load dynamics for molnupiravir, an anti-viral intervention with a different mechanism of action than the nAb cocktails. The model matches the trajectories of the change in viral load from baseline for both placebo and treatment arms [Supplementary Figure S5].

Subsequently, based on the use of the plasma IL-6 threshold of 40 pg/mL as the primary biomarker for clinical endpoints in the model, the simulated Blaze-1 and REGEN-COV trials are assessed for improvements in disease severity upon intervention within the virtual population. A therapeutic intervention that substantially decreases the viral load can decrease IL-6 levels and thus, the rate of simulated COVID-19 related events based on our IL-6 threshold [**Figure 5A**]. The rates of medically attended visits or hospitalization in the placebo and treatment arms of the virtual populations for the Blaze-1 Ph3 and REGEN-COV Ph2 trials are appropriately matched to the observed event rates from the corresponding clinical trials [**Figure 5B**]. Following from this, the model also captures the observed relative risk reduction in event rates. Hence, the model adequately captures the primary endpoints of the aforementioned clinical trials and provides a proof of concept for our approach to translate from key model states to disease severity [**Figure 5C**].

**Figure 5:**
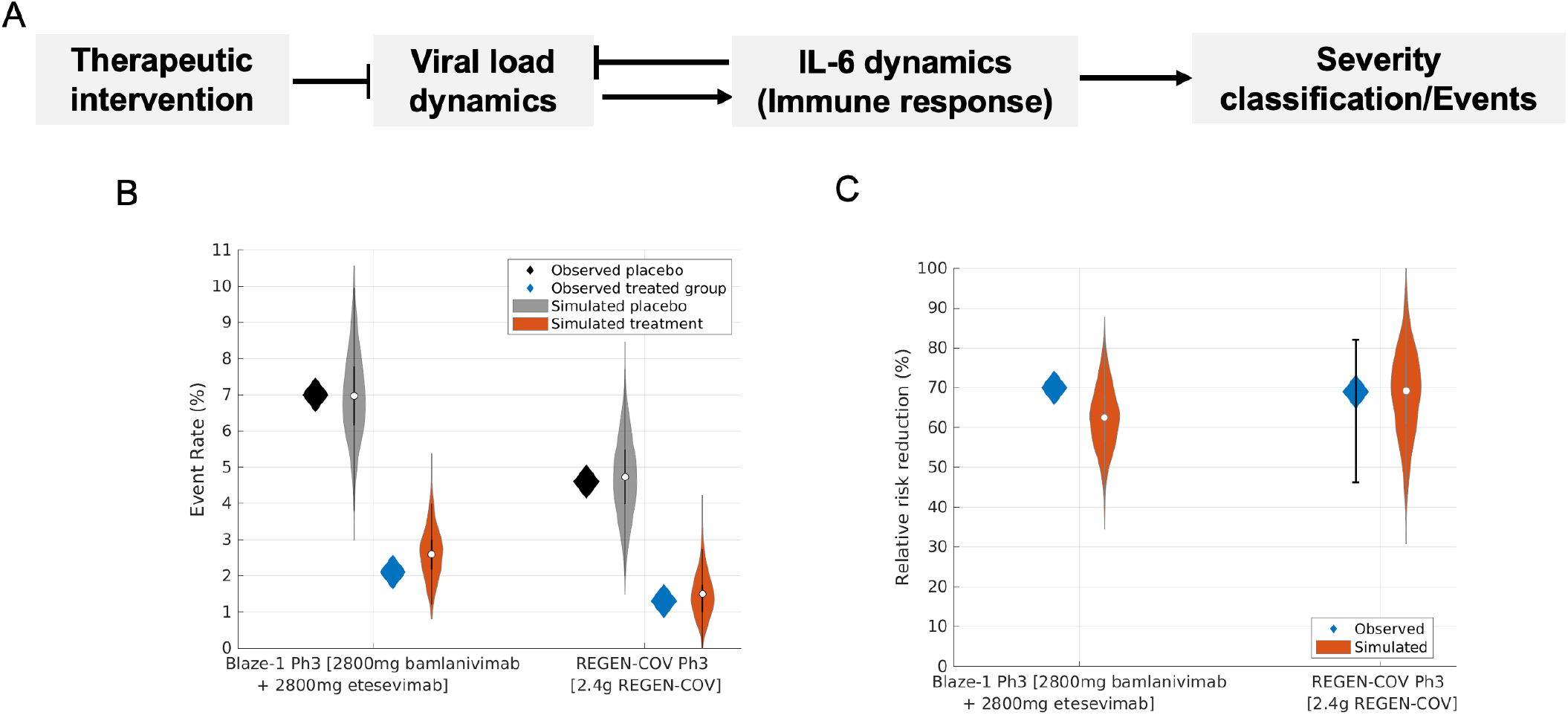
A) Workflow depicting interplay between viral load dynamics and plasma IL-6 levels, the key biomarker used to stratify COVID-19 severity. A therapeutic intervention that substantially decreases the viral load can decrease plasma IL-6 levels and thus, the rate of COVID-19 related events. B) Event rates in the observed and simulated placebo and treated group respectively for the Blaze-1 Ph3 and 2.4mg REGEN-COV Ph3 treatment arm. C) Relative risk reduction in classified events in the simulated and observed Blaze-1 Ph3 and 2.4mg REGEN-COV Ph3 treatment arms, respectively. Blaze-1 Ph3 virtual population, N=502, REGEN-COV virtual population, N=402.

Finally, we determined the sensitivity of viral load lowering efficacy and severity reduction to the time of therapeutic intervention relative to symptom onset (time of peak viral load in the model). In general, the model predicts that early intervention when closer to the time of peak viral load (symptom onset) results in greater efficacy, depicted in **Figure 6** for the Blaze-1 virtual population upon the administration of the 2800mg bamlanivimab and 2800mg etesevimab nAb cocktail. The model predicts that intervention prior to 6d post symptom onset results in greater than 50% improvement in severity outcomes, on average. Efficacy is predicted to decline rapidly as the timing of intervention is delayed to beyond 7d relative to the time of onset of symptoms. A qualitatively similar trend for the dependence of efficacy on time of intervention is also observed for simulated REGEN-COV nAb therapy [**Supplementary Figure S6**]. Despite predicting higher therapeutic efficacy at early times of intervention (<=4d) compared to later intervention (>4d), a slight non-monotonic response is predicted to occur in our virtual population simulations, when intervention occurs very early (<2d post symptom onset). This non-monotonic response occurs due to the selection of specific virtual subjects in our population, where very early intervention is found to increase the AUC of viral load post dosing compared to placebo conditions, given the PK/PD parameters of the simulated treatments. While this subset of patients can be filtered from the final virtual population as further appropriate real-world clinical data becomes available, given the current relative paucity of outpatient data, it is possible that these virtual subjects are representative of a model-identified risk where earlier intervention might not always lead to substantially greater clinical benefit.

**Figure 6:**
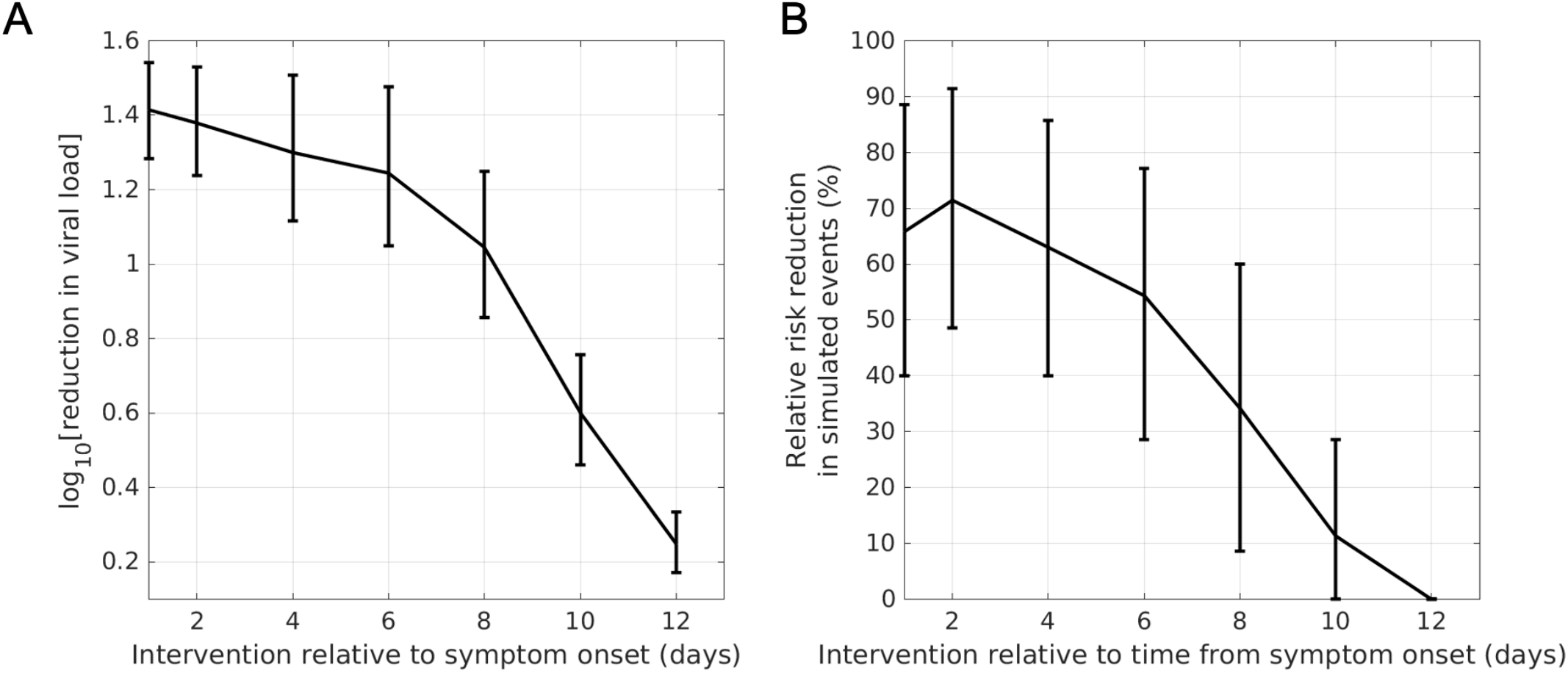
Sensitivity of A) viral load lowering efficacy and B) disease severity reduction to the time of intervention for the simulated 2800mg bamlanivimab + 2800mg etesevimab treatment and Blaze-1 Ph3 virtual population. The model predicts that early intervention when closer to peak viral load (symptom onset) results in greater viral load lowering efficacy and relative risk reduction in severity end-point. Error bars indicate 99% prediction intervals of mean.

## Discussion

To adequately inform drug development and clinical trial design decisions, QSP models must reasonably encapsulate key features of disease pathophysiology as well as appropriately represent the observed heterogeneity in real-world clinical populations. We report an updated version of our prototype model of COVID-19 with a robust virtual population capable of capturing the key viral load and severity endpoints from outpatient RCTs of therapeutic interventions targeting the viral dynamics of SARS-CoV-2. Furthermore, in recapitulating both nAb and anti-viral RCTs, the model can capture clinical responses with distinct mechanisms of action.

Moreover, our results also act as a proof of concept for the relatively simple approach we employed to translate the QSP model outputs to disease severity metrics. As more mechanistic information becomes available, the QSP model lends itself to the incorporation of clinical information on additional biomarkers, such as ferritin and CRP, which are preliminarily implemented in the model [47]. More sophisticated probabilistic approaches, e.g. Markov-chain based models, might also be used to account for the inherent uncertainty in biomarker-based classification of disease severity and clinical trajectories of COVID-19 patients. Furthermore, data-driven approaches for the prognosis of COVID-19 disease progression might be leveraged for the calibration of additional biomarkers and ultimately inform QSP model-predicted improvements in disease prognosis upon therapeutic intervention [68].

The robustness of the virtual population is partially validated using independent clinical data from the REGEN-COV Ph2 and Ph3 RCTs. Given only baseline viral load information from the Ph2 trial, the model predicts the viral load trajectories of the placebo and treatment arms. Notably, as further validation, in agreement with clinical observations [24,50], the model further predicts that subjects with higher baseline viral load exhibit larger reductions in viral load upon treatment, a finding borne out solely from the dynamics of the model. These observations support the predictive potential of the QSP model and reported virtual population to inform key decisions during the development of novel therapeutic interventions.

An important factor in the deployment of effective pharmaceutical therapies and consequently, clinical trial design is identifying patient populations that will most benefit from an intervention. We find that the clinical efficacy of pharmaceutical intervention is sensitive to the timing of intervention relative to the time of symptom onset. The model predicts that intervention within 6 days relative to symptom onset on average would be necessary to achieve meaningful clinical efficacy in outpatients with mild to moderate COVID-19 severity. Our predictions are supported by recently published RCTs, which suggest that early intervention improves clinical outcomes in this patient population. RCTs in COVID-19 outpatients limiting recruitment of subjects to within 5-7 days post symptom onset have shown clinically meaningful improvements in clinical outcomes [22,50,69,70]. Furthermore, both nAbs and anti-viral therapies were found to be at most marginally effective in reducing mortality in hospitalized, COVID-19 patients, leading to discontinuation of larger clinical trials in these patients due to low likelihood of benefit [69,71-73]. Albeit a different population than the one studied here, the reduced efficacy of virus targeting treatments in hospitalized patients is likely contributed by the fact that such patients are further along the disease course, with the reported average time of symptom onset to hospitalization being 8-10 days [42,49,74]. The model predicts that at later times in the disease course, the immune response will likely contribute more to disease pathology with viral loads having decreased by several orders of magnitude relative to peak viral loads. More recently, early read-outs from a clinical trial of AZD7442, a long-acting nAb combination more closely analyzed the sensitivity to timing of intervention in a prespecified analysis in COVID-19 outpatients enrolled within 7 days of symptom onset. While hospitalization rates decreased by 50% for the overall trial population, patients treated within 5d of symptom onset exhibited a 67% decrease in risk of hospitalization [75]. These observations are in remarkably close alignment with our predictions and lend further credence to the predictive utility of the model in informing key clinical trial design parameters, such as inclusion criteria.

In qualitative agreement with our results, previous systems modeling studies also find that early intervention post infection is required for adequate therapeutic efficacy [15,16,76]. However, many prior models predicted that viral intervention post peak viral load or more than 1-2 days post symptom onset would likely not result in clinical efficacy [77]. Supporting our assumption that symptom onset occurs at peak viral load, COVID-19 outpatients in recent RCTs, enrolled on average within 4-5 days of symptom onset are already post peak viral load. Our model suggests a relatively slow attenuation of efficacy with meaningful reductions in the risk of hospitalization predicted to occur with interventions starting up to 5d post peak viral load or symptom onset. The less pronounced attenuation of efficacy with time from peak viral load can be at least partially attributed to the log-sensing activation of the immune response in the QSP model, thus enabling the immune system to be comparably responsive as the viral antigen varies over orders of magnitude. Therefore, the model suggests that RCTs in COVID-19 outpatients might preferentially recruit patients within 5d post symptom onset to appropriately evaluate the efficacy of therapeutics.

The model lends itself to several potential additional analyses not presented in this work. In this study, we focused on developing a robust virtual population to support the development of therapeutic interventions applicable to COVID-19 outpatients. However, given the generality of our tiered approach and comprehensive mechanistic architecture of the model, the model can be used to develop robust virtual populations in other patient populations, as in hospitalized patients with severe COVID-19. While the focus of the current study has been on modeling anti-viral interventions, given the comprehensive immune component of the model, additional interventions of interest especially immunomodulatory interventions can be subsequently incorporated and validated against emerging clinical trial data in hospitalized COVID-19 patients. Moreover, the model can be adapted to multiple therapeutic scenarios, such as pre- and post-exposure prophylaxis. This is especially relevant to the treatment of close-contacts of infection-confirmed cases, with a number of clinical trials exploring the efficacy of pharmaceutical interventions in such settings [78]. Furthermore, virtual populations might also be constructed to match viral load and immune dynamics in vaccinated individuals upon breakthrough infection as more RCT or other appropriate datasets on such subjects become available.

The model calibration procedure described in preceding sections can be used to obtain a virtual population representative of SARS-CoV-2 variants that might exhibit differing viral dynamics compared to the SARS-CoV-2 clades prevalent in the 2019-2021 period of the COVID-19 pandemic, including the Alpha and Beta variants. We developed a preliminary, proof-of-concept virtual population matching the virological characteristics of the Delta variant, the dominant variant of concern worldwide as of Nov 2021. While we do not use the model to predict the effects of pharmaceutical interventions in the Delta variant infections given the current substantial uncertainty in the differences in disease pathophysiology, and viral dynamics between the Delta variant and older variants of SARS-CoV-2, as more data becomes available the preliminary virtual population presented in this work can be adapted to address such questions [Supplementary Figure S7]. Emerging SARS-CoV-2 variants of concern can potentially impact the epidemiological properties of COVID-19, such as to changes in infectiousness, associated disease severity. Variants of concern can warrant a re-consideration of the efficacy of pharmaceutical interventions. For instance, despite considerable efficacy against the 2019-2020 clades of SARS-CoV-2, the FDA withdrew the EUA for the use of bamlanivimab alone in 2021 due to evidence showing significantly reduced efficacy against the Delta variant, which was quickly becoming the dominant variant of concern in the United States [79,80]. Given these potential implications, mechanistic model-informed analysis can help address questions associated with how variants can impact key drug development parameters, including changes to the dose/dosing regimen, the development of new anti-viral combinations and the withdrawal altogether of therapies no longer effective against more recent variants.

In summary, the QSP model is to our knowledge, currently the only model capable of quantitatively capturing key clinical end-points from recently conducted interventional RCTs in outpatient populations involving therapies with distinct mechanisms of action. We presented a robust virtual population, which was partially validated against the REGEN-COV RCT and is capable of informing key clinical trial design parameters for novel COVID-19 interventions. There are number of limitations to our approach. Chiefly, while model components directly describing the viral load dynamics are calibrated against both hospitalized and outpatient datasets, the majority of immune states are informed by data in hospitalized COVID-19 subjects. Additionally, the model does not distinguish between distinct compartments of infection, such as the upper and lower respiratory tract, and further does not account for the mechanistic influence of excessive immune activation on the incidence of systemic complications or the impact of systemic comorbidities on disease severity. Finally, we do not comprehensively account for the endogenous humoral SARS-CoV-2 antibody response dynamics, which is found to be associated with baseline viral load. Subsequent releases of the model will focus on addressing these limitations and extending the model to other patient-care settings, such as in the case of high-risk vaccinated subjects with pre-existing immunity and the development of immunomodulatory treatments in hospitalized patients.

## Supporting information

Supplementary Equations

## Data Availability

All model files and simulations produced are available online at https://github.com/openPfizer/QSP_model_COVID19

https://doi.org/10.5281/zenodo.5745883

## Acknowledgements

We sincerely thank Annaliesa Anderson, Arthur Bergman, Britton Boras, Phylinda Chan, Wei Dai, Bharat Damle, Sandeep Menon, Gianluca Nucci, Theodore Rieger, Ravi Singh, Nessy Tania and, RES group for their comments and feedback on the manuscript and during the development of the model.

