## Supplementary Figures & Tables for "A Quantitative Systems Pharmacology Model of the Pathophysiology and Treatment of COVID-19 Predicts Optimal Timing of Pharmacological Interventions"

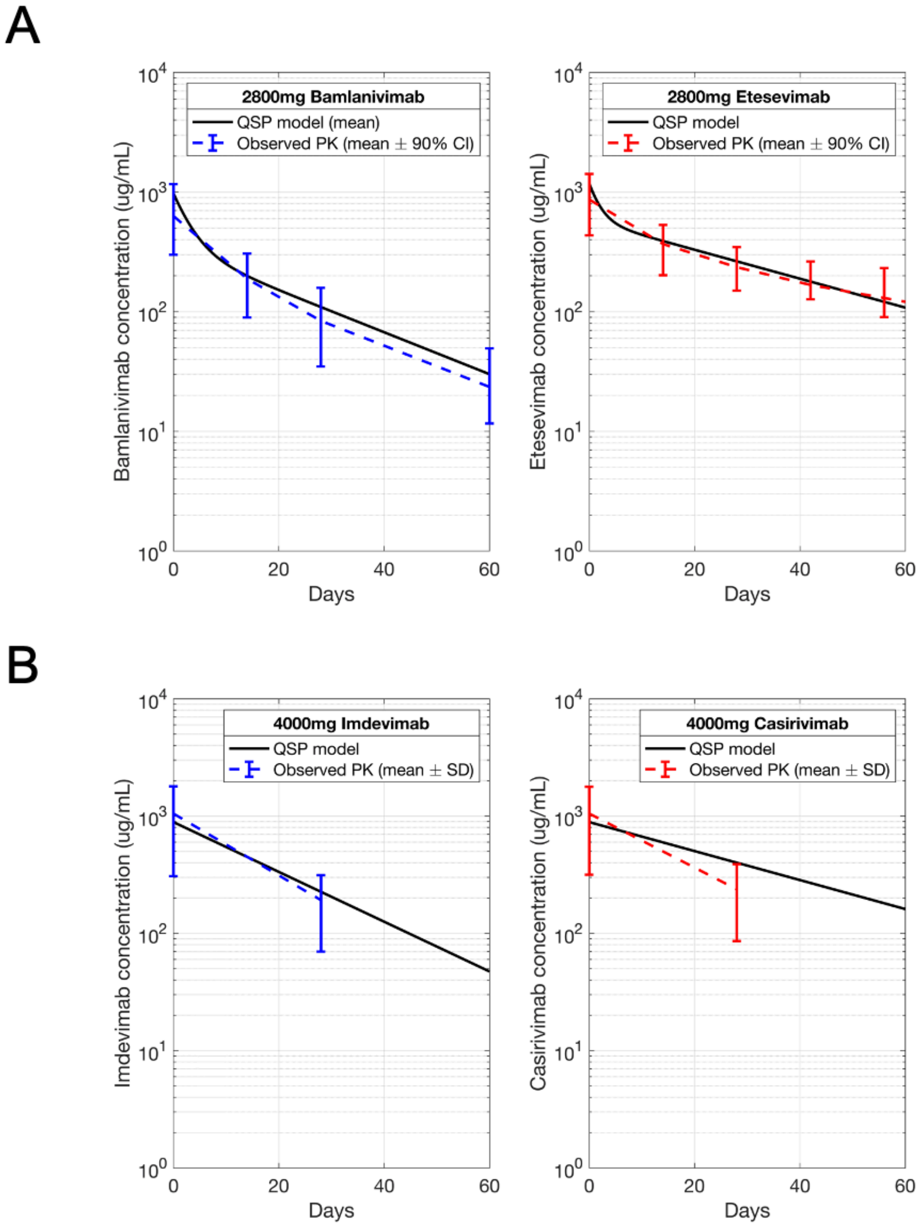


Figure S1: Observed and simulated plasma concentration profiles for A) 2800mg bamlanivimab and 2800mg etesevimab from the Blaze-1 Ph3 trial B) 4000mg imdevimab and 4000mg casirivimab from the REGEN-COV Ph2 trial.

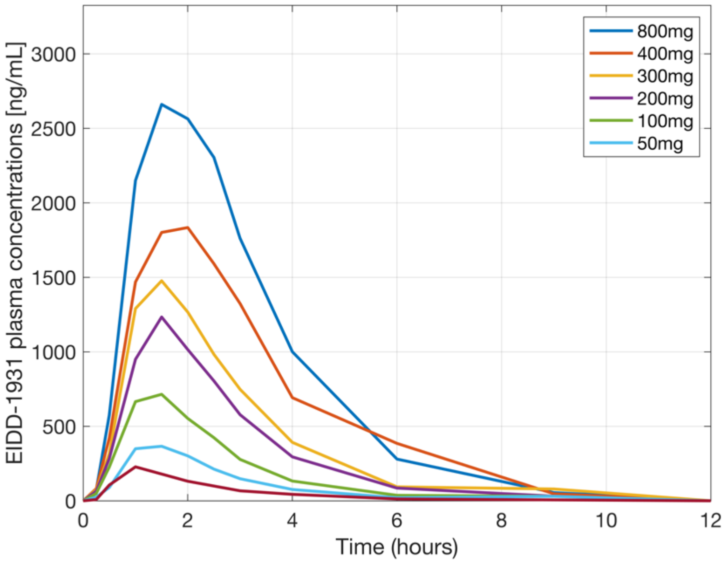


Figure S2: Plasma concentration profile of EIDD-1931 (active metabolite of molnupiravir) extracted from Painter et al. [1].

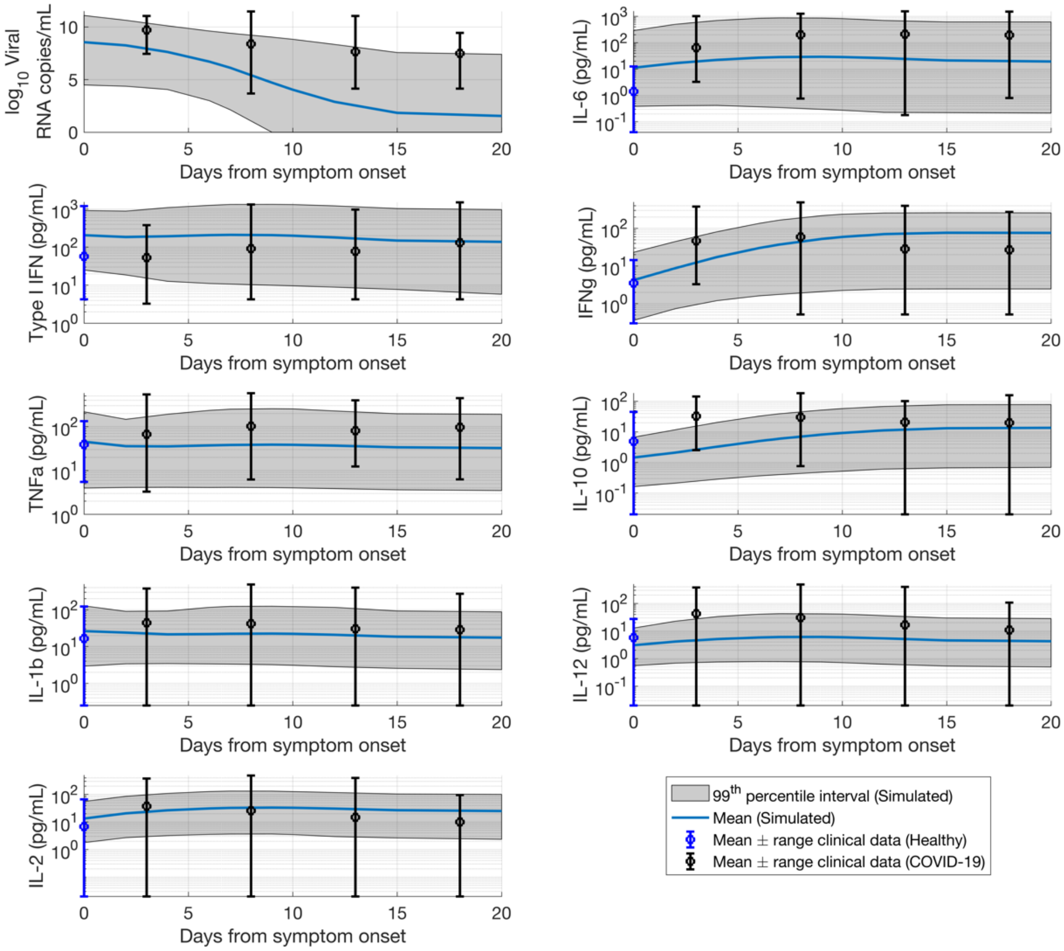


Figure S3: Plausible population (N = 14545) overlaid against observational COVID-19 clinical data for the viral load time course and different representative cytokines. The time course is presented relative to days from symptom onset given the assumption the symptom onset coincides with viral load peak for each virtual subject.

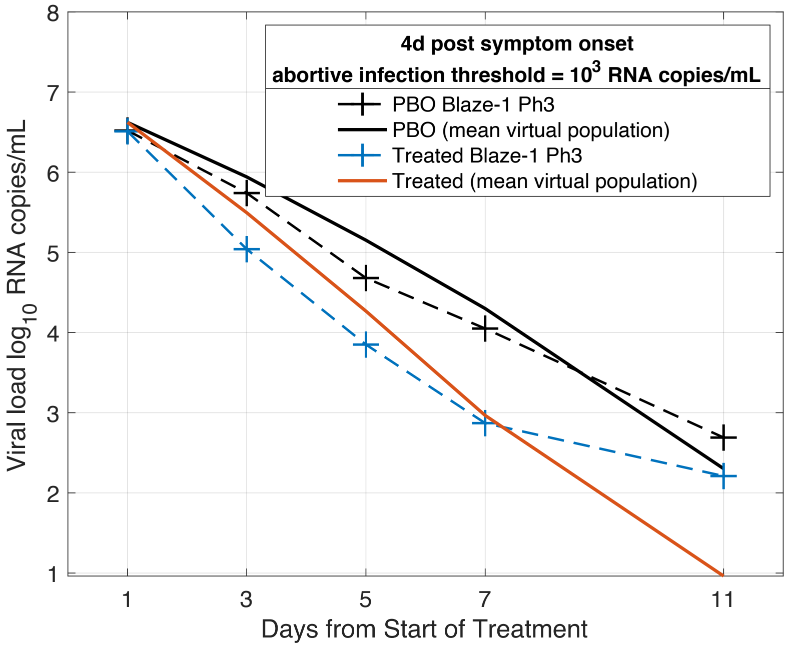


Figure S4: Virtual population matching the observations from the Blaze-1 Ph3 trial where the threshold for inactive virus is 1000 RNA copies/mL. Mean of the virtual population (N=502) for the simulated placebo (PBO) group and the 2800mg bamlanivimab + 2800mg etesevimab simulated treated group matching the mean trial data from the observed Blaze-1 Ph3 placebo group and the 2800mg bamlanivimab + 2800mg etesevimab treated group.

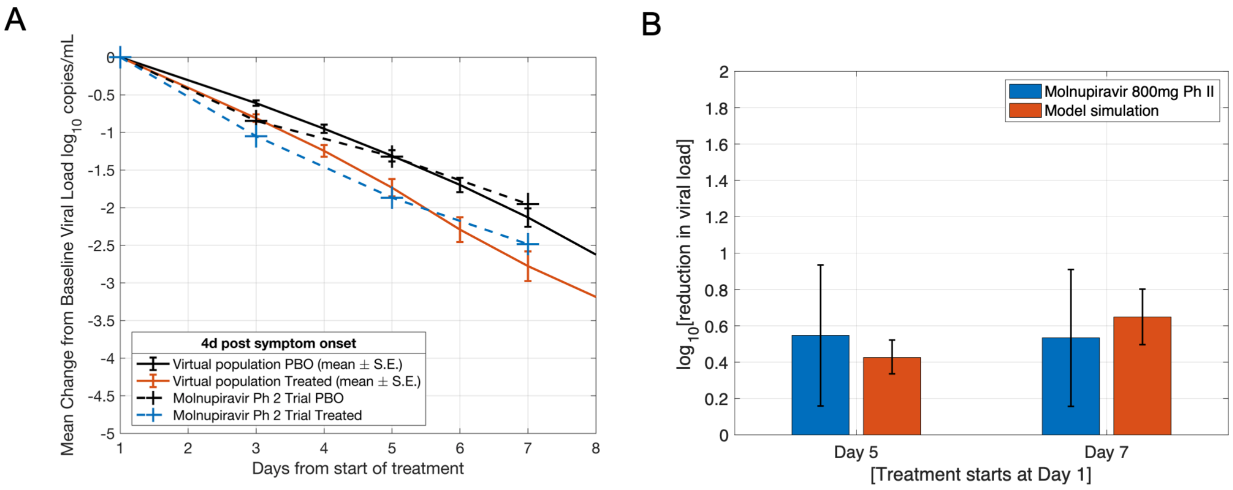


Figure S5: Virtual population matched to recapitulate A) the observed time course of the mean change in viral load from baseline for the placebo and treated groups upon administration of 800mg Molnupiravir BID, B) the reduction in viral load from baseline at Day 5 and Day 7 from treatment.

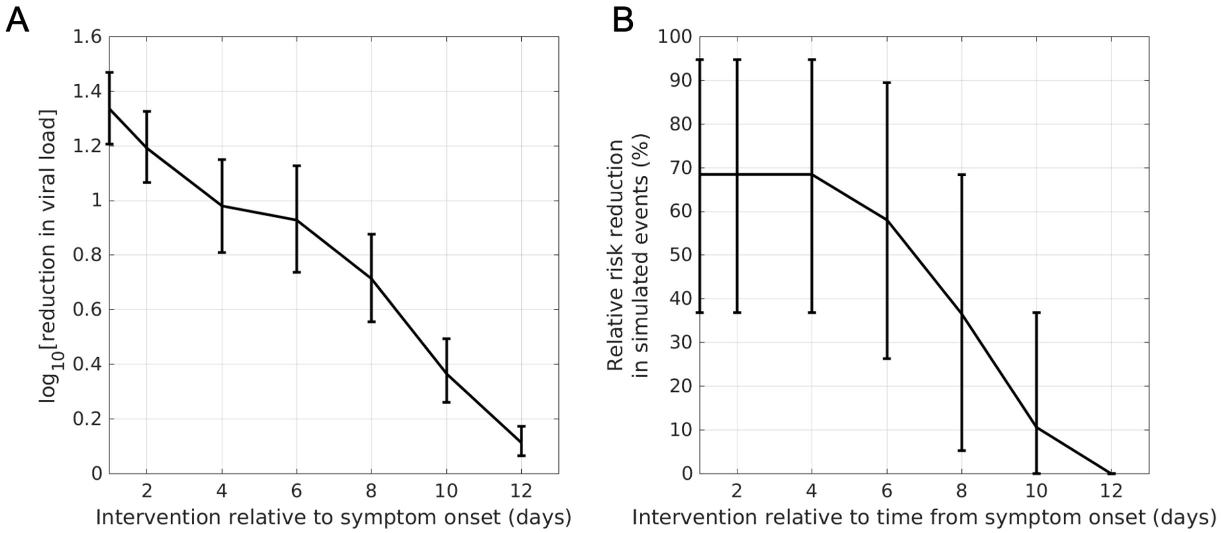


Figure S6: A) Virological and B) clinical efficacy exhibiting similar sensitivity to the time of intervention after treatment with REGEN-COV nAb cocktail.

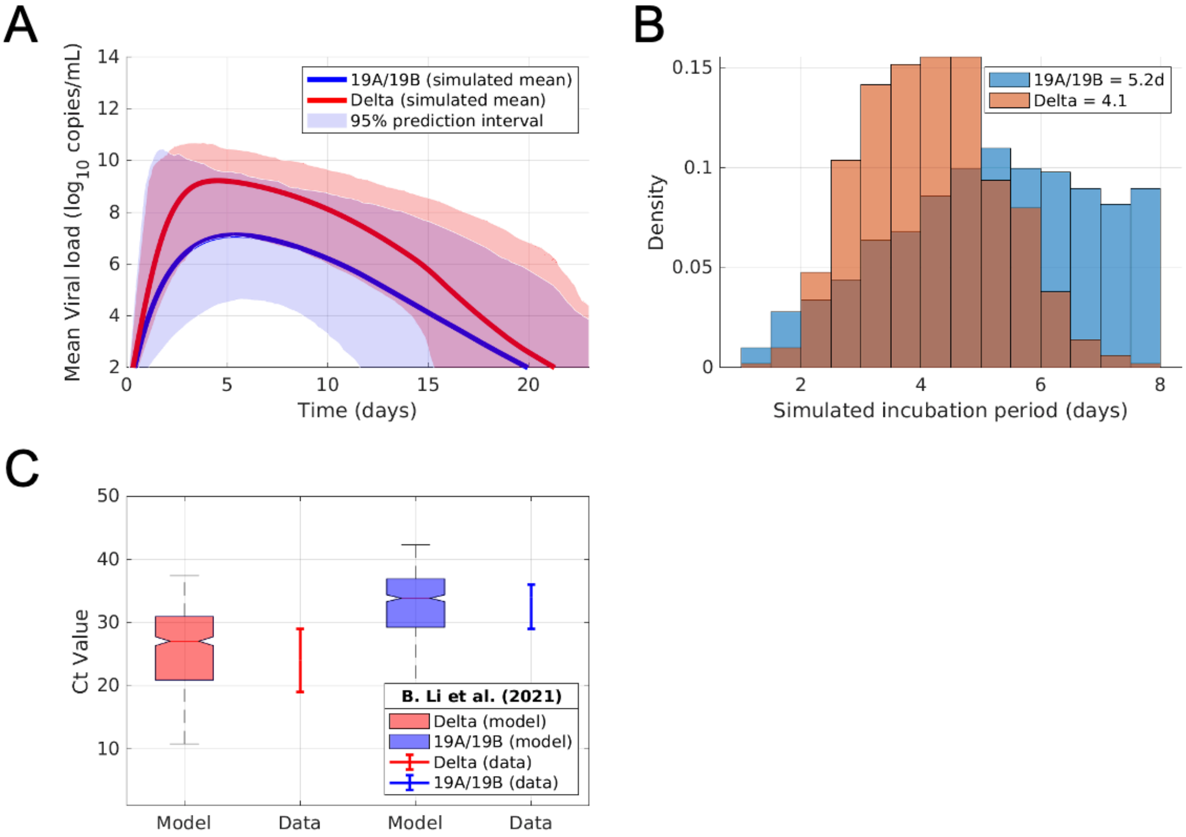


Figure S7: Virtual population with delta variant viral dynamics. A) Li et al. provided viral load measurements enabling an approximate conversion between PCR assay Ct values and RNA copies/mL. Virtual subjects were selected from the previously developed plausible population to match the reported viral load measurements in Li et al, assuming the viral dynamics from the RCT-matched virtual population were equivalent to the viral dynamics of the reported 19A/19B clade and assuming the PCRs assays were performed ~2d post infection for both the 19A/19B clade and Delta variant. B) The simulated incubation period for the Delta variant and 19A/19B clade virtual populations C) The simulated viral load time course of the Delta variant virtual population (red) compared to the non-delta SARS-CoV-2 clades prevalent in 2019-2020.

### Supplementary Table S1: Datasets used to construct plausible population

| | References |
|---|---|
| 1 | Lucas, C.; Wong, P.; Klein, J.; Castro, T.B.R.; Silva, J.; Sundaram, M.; Ellingson, M.K.; Mao, T.; Oh, J.E.; Israelow, B., et al. Longitudinal analyses reveal immunological misfiring in severe COVID-19. *Nature* **2020**, *584*, 463-469, doi:10.1038/s41586-020-2588-y. |
| 2 | Mudd, P.A.; Crawford, J.C.; Turner, J.S.; Souquette, A.; Reynolds, D.; Bender, D.; Bosanquet, J.P.; Anand, N.J.; Striker, D.A.; Martin, R.S., et al. Distinct inflammatory profiles distinguish COVID-19 from influenza with limited contributions from cytokine storm. *Sci Adv* **2020**, *6*, doi:10.1126/sciadv.abe3024. |
| 3 | Mann, E.R.; Menon, M.; Knight, S.B.; Konkel, J.E.; Jagger, C.; Shaw, T.N.; Krishnan, S.; Rattray, M.; Ustianowski, A.; Bakerly, N.D., et al. Longitudinal immune profiling reveals key myeloid signatures associated with COVID-19. *Sci Immunol* **2020**, *5*, doi:10.1126/sciimmunol.abd6197. |
| 4 | Gastine, S.; Pang, J.; Boshier, F.A.; Carter, S.J.; Lonsdale, D.O.; Cortina-Borja, M.; Hung, I.F.; Breuer, J.; Kloprogge, F.; Standing, J.F. Systematic review and patient-level meta-analysis of SARS-CoV-2 viral dynamics to model response to antiviral therapies. *medRxiv* **2020**. |

### Supplementary Table S2: Parameters varied to generate plausible population

| Parameter | Description |
|---|---|
| A_V | viral shedding by infected cells |
| b_V | endogenous viral clearance |
| b_I | death rate for infected cells |
| a_DC | rate constant for production of mature dendritic cells |
| km_DC_IL10 | IC50 for inhibition of DC activation by IL-10 |
| a_Th1 | rate constant for activation of Th1 cells |
| a_Th17 | rate constant for activation of Th17 cells |
| a_Treg | rate constant for Treg activation |
| a_M1 | rate constant for activation of macrophages |
| a_CTL | rate constant for CTL activation |
| a_ifnb | basal induction of Type I IFN |
| b_dAT1 | death rate for damaged AT1 cells |
| km_int_IFNb | IC50 for anti-viral effects of Type I IFN |
| k_v | rate constant for viral activation of innate immune cells |
| k_I | rate constant for innate immune activation by infected cells |
| k_dAT | rate constant for innate immune activation by damaged cells |
| k_kill | rate constant for infected cell clearance by CD8+ cell clearance |
| k_damage_cyt | rate constant overall cytokine damage |
| k_int | viral endocytosis by AT2 |
| basal_tnfa | basal production rate of TNF |
| basalil6 | basal production rate of IL-6 |
| basalil1 | basal production rate of IL-1 |
| basalifng | basal production rate of IFNg |
| basalifnb | basal production rate of Type I IFN |
| basalil2 | basal production rate of IL-2 |
| basalil12 | basal production of IL-12 |
| basalgmcsf | basal production of GM-CSF |
| basalil10 | basal production rate of IL-10 |
