## Supplementary Equations for "A Quantitative Systems Pharmacology Model of the Pathophysiology and Treatment of COVID-19 Predicts Optimal Timing of Pharmacological Interventions"

December 2, 2021

### 2-compartment PK model for bamlanivimab and etesevimab

$$\frac{dAb_{1c}}{dt} = -Ab_{1c} \cdot \frac{CL_{Ab1c}}{V_{Ab1c}} - Q_a \cdot \frac{Ab_{1c}}{V_{Ab1c}} + Qa \cdot \frac{Ab_{1p}}{V_{Ab1p}}$$

$$\frac{dAb_{1p}}{dt} = Q_a \cdot \frac{Ab_{1c}}{V_{Ab1c}} - Q_a \cdot \frac{Ab_{1p}}{V_{Ab1p}}$$

$$\frac{dAb_{2c}}{dt} = -Ab_{2c} \cdot \frac{CL_{Ab2c}}{V_{Ab2c}} - Q_b \cdot \frac{Ab_{2c}}{V_{Ab2c}} + Q_b \cdot \frac{Ab_{2p}}{V_{Ab2p}}$$

$$\frac{dAb_{2p}}{dt} = Q_b \cdot \frac{Ab_{2c}}{V_{Ab2c}} - Q_b \cdot \frac{V_{Ab2p}}{Ab_{2p}}$$

### Equations describing phenomenological logarithmic activation of innate immune cells

### 1 Dendritic Cell Matuaration $(DC)$

$$\frac{dDC}{dt} = \alpha_{DC}\left[k_V \cdot log(V) + k_I.log(I) + k_D log(DAT1 + DAT2)\right]\left[k_{DC(TNF)}\cdot\left(\frac{(TNF\alpha)}{K_{DC(TNF)} + (TNF\alpha)}\right) + k_{DC(IFN\gamma)}\cdot\left(\frac{(IFN\gamma)}{K_{DC(IFN\gamma)} + (IFN\gamma)}\right) + k_{DC(GM-CSF)}\cdot)\left(\frac{(GM-CSF)}{K_{DC(GM-CSF)} + (GM-CSF)}\right)\right]\left[\frac{K_{DC(IL10)}}{K_{DC(IL10)} + (IL10)}\right] - \beta_{DC}.(DC)$$

Mature $(DC)$ are formed upon recognition of viral particles $(V)$, infected cells $(I)$, and damaged $(AT1)$ and $(AT2)$ cells. $(DC)$ maturation is further induced by $(TNF\alpha)$, $(IFN\gamma)$, $(GM-CSF)$, and inhibited by $(IL10)$. Mature $(DC)$ undergo nonspecific clearance at at a rate $\beta_{DC}$.

### 2 Macrophage Activation $(M1)$

$$\frac{dM1}{dt} = \alpha_{M1}\Bigg[k_V \cdot log(V) + k_I.log(I) + k_D log(DAT1 + DAT2)\Bigg]\Bigg[k_{M1(TNF)}\cdot\left(\frac{(TNF\alpha)}{K_{M1(TNF)} + (TNF\alpha)}\right) +$$

$$k_{M1(IFN\gamma)}\cdot\left(\frac{(IFN\gamma)}{K_{DC(IFN\gamma)} + (IFN\gamma)}\right) + k_{M1(GM-CSF)}.)\left(\frac{(GM-CSF)}{K_{M1(GM-CSF)} + (GM-CSF)}\right)\Bigg]$$

$$\left[\frac{K_{M1(IL10)}}{K_{M1(IL10)} + (IL10)}\right] - \beta_{M1}.(M1)$$

Activated $(M1)$ are formed upon recognition of viral particles $(V)$, infected cells $(I)$, and damaged $(AT1)$ and $(AT2)$ cells. $(M1)$ activation is induced by $(TNF\alpha)$, $(IFN\gamma)$, $(GM-CSF)$, and inhibited by $(IL10)$. Activated $(M1)$ undergo nonspecific clearance at at a rate $\beta_{M1}$.

### 3 Neutrophil Activation $(N)$

$$\frac{dN}{dt} = \alpha_{N}\Bigg[k_V \cdot log(V) + k_I.log(I) + k_D log(DAT1 + DAT2)\Bigg]\Bigg[k_{M1(TNF)}\cdot\left(\frac{(TNF\alpha)}{K_{M1(TNF)} + (TNF\alpha)}\right) +$$

$$k_{M1(IFN\gamma)}\cdot\left(\frac{(IFN\gamma)}{K_{DC(IFN\gamma)} + (IFN\gamma)}\right) + k_{M1(GM-CSF)}.)\left(\frac{(GM-CSF)}{K_{M1(GM-CSF)} + (GM-CSF)}\right)\Bigg]$$

$$ktr_N \cdot N_c \cdot (\frac{(IL17)}{km_{N\_IL17} + (IL17)}) - \beta_N \cdot N - k_{tr(N)} \cdot N$$

Activated $(N)$ are formed upon recognition of viral particles $(V)$, infected cells $(I)$, and damaged $(AT1)$ and $(AT2)$ cells. $(N)$ activation is induced by $(TNF\alpha)$, $(IFN\gamma)$, $(GM-CSF)$, and inhibited by $(IL10)$. Activated $(N)$ undergo nonspecific clearance at at a rate $\beta_N$. Activated $(N)$ migration is also induced by $(IL17)$.
